# Integrated Histopathology–Transcriptomic Biomarker Enhances Survival Prediction in HNSCC Patients Treated with Immunotherapy

**DOI:** 10.1101/2025.11.26.25341072

**Authors:** Merzu Belete, Nitya Thakkar, Indu Khatri, Meijian Guan, Yu Sun, Anantharaman Muthuswamy, Iris Kolder, Han Si, David Soong, Brandon W. Higgs, Lauren K. Brady, James Zou, Sriram Sridhar

## Abstract

Recurrent and metastatic head and neck squamous cell carcinoma (R/M HNSCC) remains a challenging disease with modest response to immune checkpoint inhibitors and a need for more robust predictive biomarkers. In a real-world (RW) cohort of patients treated with pembrolizumab alone or in combination with chemotherapy, we evaluated transcriptomic and histopathologic features associated with therapeutic benefit. PD-L1 expression, measured by combined positive score (CPS), was not significantly associated with progression-free survival (PFS). In contrast, immune-related gene signatures, particularly those linked to T cells and tertiary lymphoid structures (TLS), were predictive of improved outcomes. TLS presence identified from Hematoxylin and Eosin-stained (H&E) whole slide images (WSI) correlated with favorable survival and showed concordance with RNA-derived TLS signatures. TLS-associated features demonstrated treatment-specific prognostic patterns, with stronger predictive power in pembrolizumab monotherapy versus combination therapy. We developed multimodal risk prediction models integrating molecular features with imaging data which better associated with RW outcomes. Evaluation using concordance index analysis revealed that traditional pathological markers and individual molecular signatures had modest predictive capability. Digital pathology features achieved better performance than clinical or molecular features alone, but the combination of imaging and molecular features yielded the highest predictive accuracy with concordance index values of 0.86 and 0.81 in pembrolizumab and combination therapy cohorts, respectively. Kaplan-Meier analysis confirmed that our multimodal risk signature achieved significant separation between high- and low-risk groups in both treatment arms, substantially outperforming molecular features alone. These findings highlight that integrating transcriptomic and histopathological data enables precise patient stratification for immunotherapy in R/M HNSCC.

**Significance:** Multimodal risk signatures combining transcriptomic and imaging data significantly outperform individual biomarkers including PD-L1 and TLS in predicting immunotherapy response, enabling superior patient stratification in R/M HNSCC.

## Introduction

Head and neck squamous cell carcinoma (HNSCC) constitutes a diverse group of epithelial malignancies that arise from the mucosal surfaces of the oral cavity, pharynx, and larynx, and represents one of the most prevalent cancer types globally [1]. HNSCC represents approximately 4.5% of all malignancies worldwide, with an estimated 890,000 new cases annually, ranking it as the seventh most common cancer type. It also accounts for roughly 4.5% of global cancer-related mortality (450,000 deaths per year) [2]. Notably, the incidence of HNSCC has been rising, with a disproportionate increase observed among younger patient populations [3].

HNSCC is biologically and clinically heterogeneous, encompassing tumors with distinct anatomical sites, etiological factors, and molecular characteristics. Despite advances in diagnostics and therapy, outcomes remain poor for a significant proportion of patients, particularly those presenting with locally advanced or recurrent/metastatic (R/M) disease [4].

Most patients with HNSCC are diagnosed at a late stage, often in the absence of recognizable pre-malignant lesions. The presence or absence of human papillomavirus (HPV) has emerged as a critical prognostic biomarker, especially in oropharyngeal cancers. HPV-positive tumors are associated with better prognosis and increased responsiveness to therapy [5] while HPV-negative tumors tend to exhibit more aggressive behavior and therapeutic resistance [6, 7].

The current treatment landscape for HNSCC is challenged by multiple factors. Conventional therapies in the locally advanced setting, including platinum-based chemotherapy, radiation, and surgery, are often associated with substantial toxicity and limited long-term efficacy [8]. Patients with R/M HNSCC have fewer treatment options, high resistance to therapy, and poor survival rates, with median overall survival (OS) ranging from 6-15 months [9]. Moreover, the genetic heterogeneity of HNSCC tumors, together with variability in their tumor microenvironment, complicates therapeutic targeting and contributes to inconsistent treatment responses. A major unmet need is the development of robust predictive biomarkers that can guide treatment selection and patient stratification [10].

Recent advances in molecular and immunotherapeutic strategies have opened new avenues for improving outcomes in HNSCC. These include the development of targeted agents, antibody-drug conjugates, and immune checkpoint inhibitors, as well as the rational design of combination therapies. Pembrolizumab is being used to treat R/M HNSCC patients as a monotherapy or in combination with chemotherapy in the 1L setting [11] while other bispecific antibody and dual-targeting therapies are emerging [12]. Given these advances, there is growing interest in applying biomarker-guided treatment strategies aimed at maximizing efficacy while minimizing toxicity. Treatment combinations incorporating immunotherapy, targeted therapy, and chemotherapy in sequential or concurrent fashion may offer a way to overcome resistance mechanisms and broaden therapeutic benefit [5, 13]. Beyond therapeutic combinations, the integration of multiple data modalities, including transcriptomic profiling, digital histopathology, and clinical features represents an emerging paradigm for developing better models for predicting therapeutic response and progression. Traditional single-biomarker approaches, such as PD-L1 expression assessed through CPS [14], have significant limitations due to high interobserver variability and poor reproducibility among pathologists [15]. This highlights the need for comprehensive, multimodal biomarker strategies that can capture the complex interplay between tumor biology and immune microenvironment.

To address these challenges, we analyzed a RW cohort of 230 R/M HNSCC patients treated with Pembrolizumab (Pembro) alone or in combination with chemotherapy (Pembro+Chemo), with the goal of identifying robust molecular and imaging signatures of response that could better reflect the complexity of the tumor and its surrounding immune microenvironment compared to PD-L1 expression alone. We systematically evaluated transcriptomic signatures, histopathological features, and their combinations to identify the most predictive approaches for treatment response. Among the molecular features examined, TLS [16] signatures emerged as particularly promising biomarkers that capture immune-rich histopathological features associated with treatment response—features that can be inferred both through bulk-transcriptomic data and direct histological evaluation [16–18]. Notably, several gene expression signatures associated with TLS, as well as pathologist-annotated TLS presence, were predictive of response particularly in patients receiving Pembro monotherapy, underscoring their potential as broadly applicable biomarkers. However, these TLS-related signatures alone were not always sufficient to optimally stratify patients based on outcomes, particularly those treated with combination immunochemotherapy. We subsequently developed and validated integrated risk prediction models that combined transcriptomic features with imaging-derived characteristics extracted from whole-slide histopathological images using deep learning approaches. This multimodal strategy was designed to capture complementary aspects of tumor biology and spatial organization that are not accessible through any single data modality. Our systematic evaluation demonstrated that while individual molecular signatures and digital pathology features showed modest predictive performance, their integration yielded substantially improved prognostic accuracy, achieving superior patient stratification across both treatment cohorts. These findings support the clinical potential of multimodal biomarker strategies for precision medicine in HNSCC immunotherapy.

## Material and Methods

### Cohort and metadata description

We examined de-identified RW patients’ records with a primary diagnosis of R/M HNSCC treated in Standard-Of-Care between June 2011 and September 2023 acquired from Tempus (Tempus AI, Inc., Chicago, IL). In this real-world study, PFS was used as a practical endpoint because it can be measured earlier and more consistently than overall survival. Real-world PFS reflects disease progression or death as documented in clinical practice and serves as a feasible, clinically meaningful surrogate for assessing treatment effectiveness in real-world settings. The dataset comprised 106 Pembro treated, and 124 Pembro+Chemo treated R/M HNSCC, where more females and younger patients were treated in Pembro+Chemo group. Biopsy samples, obtained pre-treatment, were annotated using directly assigned patient-level timelines based on treatment history. When only patient-level clinical metadata was available, corresponding biopsy annotations were inferred based on the temporally closest to clinical events within one year prior to first therapy. The majority of patients (∼97%) presented with advanced-stage disease (Stage II or IV) as detailed in patient characteristics summary Table 1. PD-L1 expression was categorized based on CPS [14] into three groups: CPS < 1 (negative), 1 ≤ CPS < 20 (low), and CPS ≥ 20 (high), with majority of the patients in the CPS-low group. Similar to clinical studies KEYNOTE-048 and KEYNOTE-B10 [13, 19], we found no significant PFS advantage between the treatment groups (Supplementary Figure S1). HPV status was known for 55 patients; for the remaining samples, a high-confidence HPV status was imputed using a trained machine learning model (see Supplementary Methods). In our RW cohort, we found no significant difference in PFS outcome among HPV positive and negative patients when stratified across the two treatment regimens.

**Table 1:** Cohort characteristics. This table describes the RW cohorts in this study where gender and age were disproportional in the two treatment groups. More female and younger age are in the Pembro treatment. PD-L1 expression was categorized based on CPS into three groups: CPS < 1 (negative), 1 ≤ CPS < 20 (low), and CPS ≥ 20 (high).

| **Characteristic** | **Pembro N = 106<sup>1</sup>** | **Pembro+Chemo N = 124<sup>1</sup>** | **p-value<sup>2</sup>** |
| --- | --- | --- | --- |
| **SEX** |  |  | 0.007 |
| *Female* | 24 (23%) | 12 (9.7%) |  |
| *Male* | 82 (77%) | 112 (90%) |  |
| **AGE** | 65 (58, 72) | 62 (54, 67) | 0.009 |
| **Biopsy Site** |  |  | 0.4 |
| *Bronchus and Lungs* | 20 (19%) | 30 (24%) |  |
| *Head, Face, Mouth, or Neck* | 53 (50%) | 51 (41%) |  |
| *Other* | 33 (31%) | 43 (35%) |  |
| **CPS** |  |  | 0.060 |
| *High* | 10 (21%) | 17 (26%) |  |
| *Low* | 37 (79%) | 42 (65%) |  |
| *Negative* | 0 (0%) | 6 (9.2%) |  |
| *Unknown* | 59 | 59 |  |
| **HPV** |  |  | 0.3 |
| *Negative* | 45 (42%) | 61 (49%) |  |
| *Positive* | 61 (58%) | 63 (51%) |  |
| **Stage** |  |  | >0.9 |
| *Stage 1* | 0 (0%) | 1 (1.2%) |  |
| *Stage 2* | 2 (3.0%) | 1 (1.2%) |  |
| *Stage 3* | 1 (1.5%) | 2 (2.4%) |  |
| *Stage 4* | 64 (96%) | 79 (95%) |  |
| *Unknown* | 39 | 41 |  |

<sup>1</sup>n (%); Median (Q1, Q3)
<sup>2</sup>Pearson's Chi-squared test; Wilcoxon rank sum test; Fisher's exact test

TLS were identified from pre-treatment H&E-stained WSIs by visual identification of dense lymphoid aggregates with organized architecture, including distinct B- and T-cell zones, located within or adjacent to the tumor. Annotations were performed manually by a trained pathologists from Genmab following established morphological criteria. Bulk transcriptome raw count data were first normalized [20, 21], and Gene Set Variation Analysis (GSVA) [22] was performed to estimate sample-wise enrichment scores for predefined gene sets. GSVA was applied using the Gaussian kernel method to compute non-parametric enrichment scores across samples, enabling the assessment of pathway-level variation independent of phenotype labels. Six published TLS signatures [16–18] scored using GSVA were used to classify patients with high or low TLS signatures. Associations between TLS high vs. TLS low and median progressive-free survival (mPFS) were assessed in each treatment groups using a Cox-proportional hazard and Kaplan-Meier models.

#### Molecular risk prediction model

To construct treatment-specific molecular risk signatures, whole-transcriptome bulk RNA sequencing data for all patients (encompassing 20K genes), subset based on the first line therapy treatment, were split into train and test sets (70/30 split, stratified by PFS event). For each treatment group, we created a risk signature on all training patients and evaluated its performance on the held-out testing patients.

First, a Cox proportional hazards model was used to determine if a gene was a significant predictor of PFS (p < 0.05 after false discovery rate correction). Then, we used a lasso regression model with a regularization penalty of 0.1 to compress this gene list (n=100-200) to 20-30 genes. Based on median risk score, the patients in the training group were characterized as high or low risk groups. We applied these genes, coefficients, and medians to the test patients to determine what their predicted risk would be if given the treatment. Model performance was evaluated using two complementary metrics: the concordance index (c-index), which quantifies the probability of correctly ranking any two patients by risk, and Kaplan-Meier risk stratification with log-rank testing, which assesses whether predefined risk groups show statistically significant outcome differences. Both metrics are necessary for clinical utility, as c-index measures individual-level discriminative accuracy required for personalized risk prediction, while significant KM stratification confirms clinically meaningful group separation.

#### WSI risk prediction model

For all patients with H&E data and PFS outcomes (n=157), we analyzed the WSIs through a weakly supervised deep learning approach using an attention-based multiple instance learning (MIL) model [23]. Each WSI was first divided into non-overlapping image patches (pixel dimensions 224×224) using CLAM for segmentation [24]. Patch level features were extracted using PLIP, a pretrained vision-language pathology foundation model [25]. These patch features were aggregated via an attention mechanism to obtain a patient-level representation. To account for different first-line therapy treatment regimens, we encoded treatment type as a categorical variable in the model. In the multimodal setting, this patient-level imaging vector was combined with the molecular-derived risk score in late fusion. The integrated features were then passed through a multilayer perceptron [26] to predict the final risk score. The model was trained using a negative log partial likelihood loss function, standard for survival analysis, with the Adam optimizer [27] to directly optimize for PFS.

We performed 5-fold cross-validation with hyperparameter tuning across the multilayer perceptron’s hidden layer size, learning rate, and dropout rate to optimize model performance. On held-out test patients, we used the trained model to predict individual risk scores. Using the median predicted risk from the training set as a threshold, we stratified test patients into high- and low-risk groups. Statistical significance of risk stratification was evaluated using the log-rank test, with p < 0.05 after false discovery rate correction considered significant.

Finally, to interpret model predictions, we visualized attention maps by overlaying learned attention weights on the original H&E images, highlighting the tissue regions most predictive of patient outcome.

## Results

### Differential prognostic of immune signatures

PD-L1 expression was evaluated for its predictive association with PFS in this RW cohort of patients with R/M HNSCC. When assessed in patients treated with pembro alone and pembro+chemo, patients grouped by PD-L1 CPS did not show any significant differences in PFS in either treatment, underscoring the need for alternative or combinatorial biomarkers (Figure 1A). To identify additional treatment biomarkers in this RW cohort, we applied Cox models adjusted for clinical covariates (age group, sex, biopsy site, and tumor purity) to selected immunotherapy-related signatures collected from published single-cell and bulk RNA-seq studies. We identified immune-related gene signatures that were significantly associated with improved PFS in both treatment groups (p < 0.05) as shown in Figure 1B, including signatures associated with cytotoxic T cells, TLS [16], and multiple Interferon-gamma inducible signatures associated with response to checkpoint blockade [28–30]. These shared signatures likely represent core immune activation pathways that confer benefit from anti–PD-1 therapy regardless of the addition of chemotherapy. Several of the signatures associated with PFS benefit (p < 0.05) specifically in the pembro-treated group included previously published signatures associated with response to pembro and atezolizumab, regulatory T cells, as well as multiple previously published TLS signatures [31]. These suggest that certain lymphoid and regulatory T cell features, particularly TLS components and memory/regulatory T cell markers [32], may be more predictive of benefit from immunotherapy alone, where immune pressure is unopposed by chemotherapy. Signatures uniquely associated with improved PFS (p < 0.05) in the pembro+chemo group included additional interferon gamma inducible genes associated with response to anti-PD-L1 therapies, as well as NK/T cell, TLS, plasma cell, and Th2 signatures. These signatures may reflect immune activation patterns or cell populations (e.g., TH2, NKT, plasma cells) that are more predictive of response in the context of chemotherapy, possibly due to modulation of the immune microenvironment or synergy with cytotoxic effects. These shared signatures represent robust immune correlates of benefit across treatment strategies. TLS and T cell–related signatures show stronger prognostic power in monotherapy, supporting their role in identifying patients who may not need chemotherapy. Several similar signatures were also observed as having prognostic significance in pembro+chemo patients (interferon gamma, TLS), while additional immune cell type signatures (Th2, NK/T) showed significance specifically in this treatment group. Of note, all of these signatures trended in the same direction in terms of their association with PFS in both treatment groups, suggesting that of the immune signatures assessed, there was common biology associated with better prognosis in both pembro and pembro+chemo patients.

**Figure 1:**
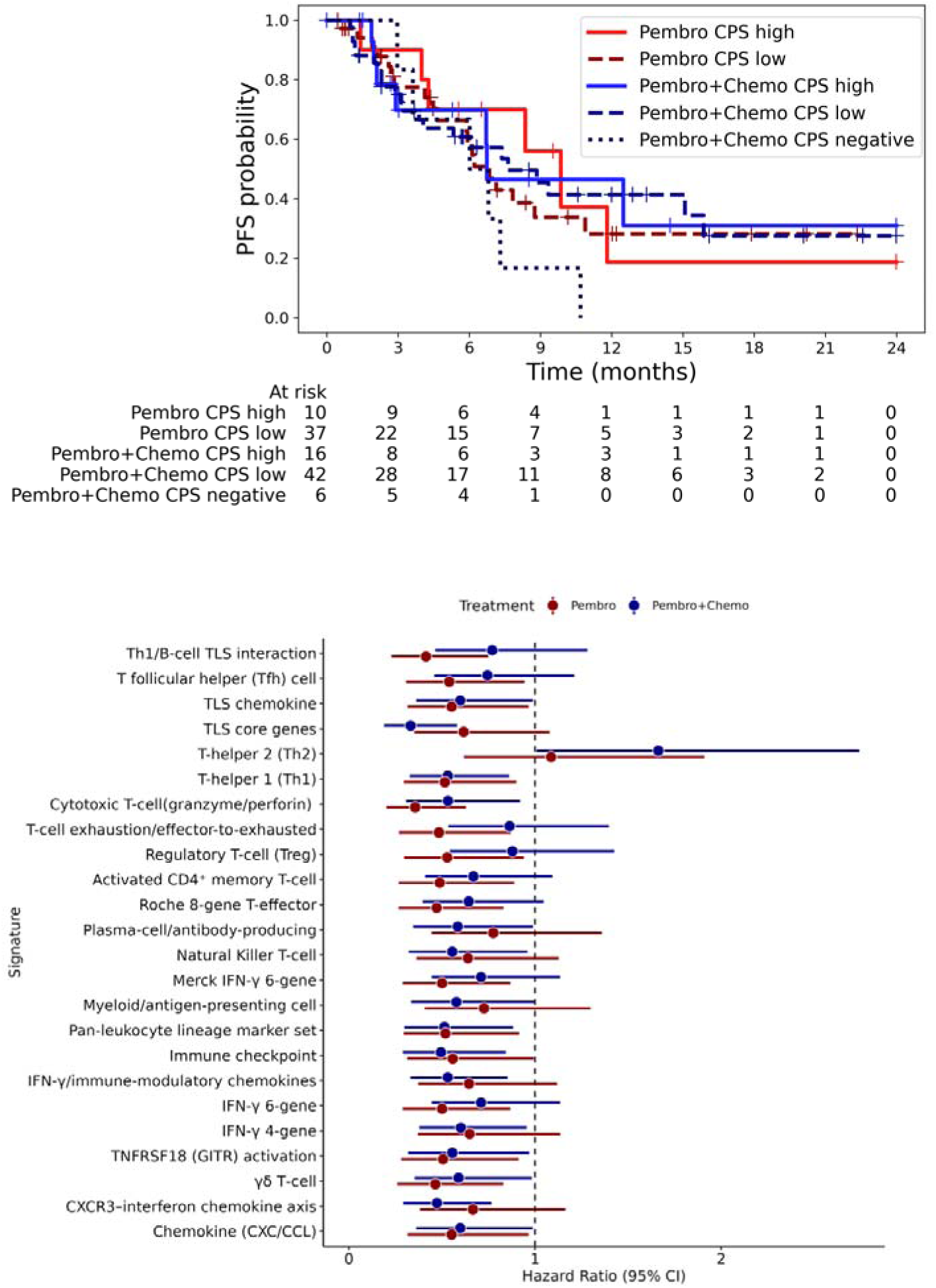
**(A)** KM plots for PFS for the treatment regimens by CPS groups . CPS did not show any significant differences in PFS in either treatment groups. **(B)** Differential prognostic of immunotherapy related signatures. We identified immune-related gene signatures that were significantly associated with improved PFS in both treatment groups.

### Subtype identifications using aggregate TLS signatures

Based on previously published associations between TLS presence and response to immunotherapy in R/M HNSCC [33], we further assessed the TLS signatures and their association with prognosis in this cohort. Most TLS-associated gene signatures demonstrated a statistically significant association with improved PFS in both Kaplan–Meier and Cox proportional hazards analyses (Supplementary Table S1). As individual TLS signatures are biased to one or other immune cell types in the TLS structures, we ran unsupervised clustering analysis using a K-nearest neighbors (KNN) approach, leveraging GSVA enrichment scores derived from six TLS gene signatures. This method enabled the stratification of samples into distinct clusters, without relying on predefined labels, and specific to a particular TLS signature. Survival analysis revealed that elevated expression of the aggregate TLS signature was associated with improved PFS, particularly in patients receiving Pembro, where the mPFS was 8.4 months in the high-expression group versus 4.8 months in the low-expression group (Figure 2A).

**Figure 2:**
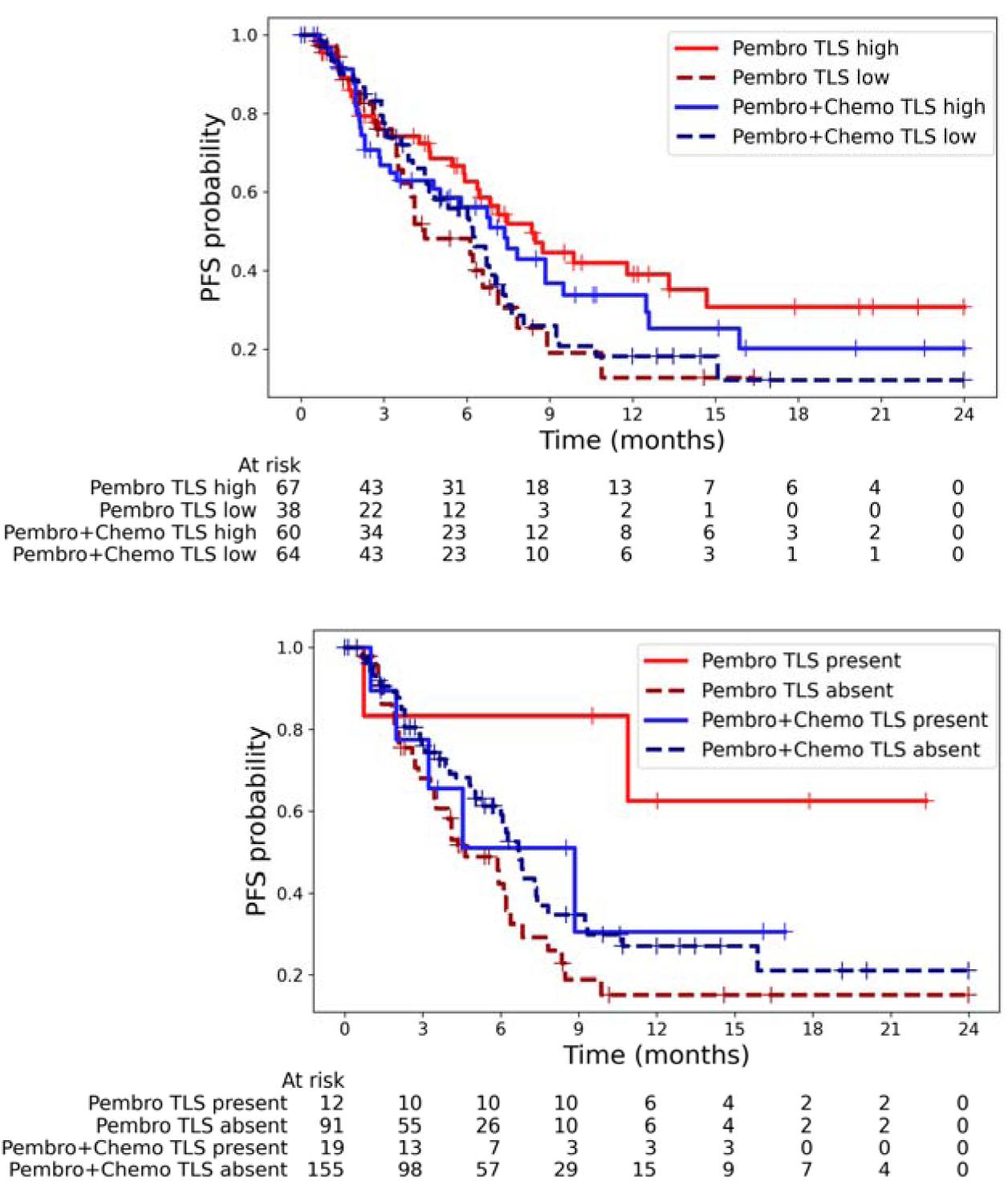
**(A)** Subtype identifications using aggregate TLS signatures. Survival analysis revealed that elevated expression of the aggregate TLS signature was associated with improved PFS, particularly in patients receiving Pembro (mPFS 8.4 vs 4.5 months) compared to Pembro+Chemo (7.4 vs 6.2 months). **(B)** TLS structure from histopathology. Presence of TLS, as determined by histopathological evaluation, was associated with improved PFS in patients receiving Pembro only (mPFS not reached -vs- 4.6 months) compared to Pembro+Chemo (8.8 vs 6.7 months).

We also identified TLS from H&E stained biopsies conducted on 277 clinical tumor samples to assess the presence (n = 31) or absence (n = 246) of TLS. The presence of TLS, as determined by histopathological evaluation, was associated with improved PFS in patients receiving Pembro monotherapy as shown in Figure 2B. Specifically, the mPFS was not reached within the 24-month follow-up period among TLS-positive patients in the Pembro group, indicating a durable clinical benefit. In contrast, a slight difference (∼ two months) in PFS was observed based on TLS status in patients treated with combination therapy consistent with result from transcriptome signature.

We next evaluated the concordance between TLS presence as determined by H&E staining from WSIs [24] and TLS-related gene expression signatures derived from bulk transcriptomic data. Five out of six TLS signatures demonstrated a statistically significant correlation with histologically defined TLS presence (p < 0.10), except for the TLS signature related to chemokine (Supplementary Figure S2 (A-F)). These findings suggest that bulk RNA-based TLS signatures can capture the presence of TLS structures observed in H&E-stained WSI, supporting the use of transcriptomic profiling as a surrogate for histopathologic TLS detection specifically in the setting of HNSCC. There is a significant enrichment of TLS-present samples in the TLS-high groups, suggesting a strong association between TLS structures and molecular groupings based on aggregate TLS-signatures (Supplementary Table S2).

We subsequently assessed the relationship between HPV status and TLS presence, as determined by both H&E-stained WSI and bulk transcriptomic data. No significant association was observed between HPV positivity and TLS presence based on histopathologic evaluation from WSI (Supplementary Table S3). However, among the transcriptomic TLS signatures, only the T follicular helper (Tfh) cell–associated signature showed a significant correlation with HPV status, suggesting a specific link between HPV-driven tumors and Tfh-related immune activity at the transcriptomic level (Supplementary Figure S3 (A-F)).

### Multimodal treatment-specific risk modeling

While individual TLS signatures showed prognostic value, we hypothesized that a data-driven approach leveraging the full transcriptome could capture additional predictive information not represented in any single literature-derived signature. We therefore developed treatment-specific molecular risk signatures to assess whether this exploratory approach could improve upon established biomarkers. For this analysis, we first compiled c-index scores for individual biomarkers, including PD-L1 CPS (based on Pathologist-annotated categorical definitions), expression-derived TLS signatures, clinical features, and whole transcriptome data using expression of individual genes (see Supplementary Tables S4, S5 for genes that comprise the molecular risk signature). We similarly evaluated the predictive performance of different feature combinations using the c-index, first training models on subsets of patient data within the Pembro and Pembro+Chemo cohorts, then applying unimodal and multimodal predictors on test patients across two treatment cohorts. We provide the exact model hyperparameters used to train these models in Supplementary Table S6.

In Figure 3A, we provide a comprehensive comparative analysis of the biomarkers’ predictive performance. In the Pembro cohort, traditional pathological markers showed limited predictive performance: the pathologist-derived CPS achieved a c-index of 0.49, while the expression-derived TLS value reached 0.43. This discordance between TLS’s significant risk stratification (Figure 2B) and poor c-index reflects the limited discriminative information in binary markers, which can separate groups but cannot accurately rank individual patients’ risk continuously. Clinical features performed similarly with a c-index of 0.54. Molecular features expanding to using whole transcriptome data as opposed to targeted signatures demonstrated improved performance at 0.66, but imaging features from WSI created using PLIP, a pre-trained pathology foundation model, showed superior predictive capability with a c-index of 0.78. The highest performance was achieved by combining imaging and molecular features, reaching a c-index of 0.86. The Pembro+Chemo cohort exhibited a comparable pattern. Traditional pathological markers again showed poor predictive value, with CPS at 0.46 and TLS at 0.41. Clinical features (c-index: 0.61), molecular features (c-index: 0.59), and imaging features (c-index: 0.57) performed at similar levels. As with the Pembro cohort, the combination of imaging and molecular features yielded the strongest predictive capability with a c-index of 0.81. These findings demonstrate that multi-modal integration of imaging and molecular features provides superior prognostic accuracy compared to traditional pathological methods, with consistent performance improvements observed across both treatment regimens.

**Figure 3.**
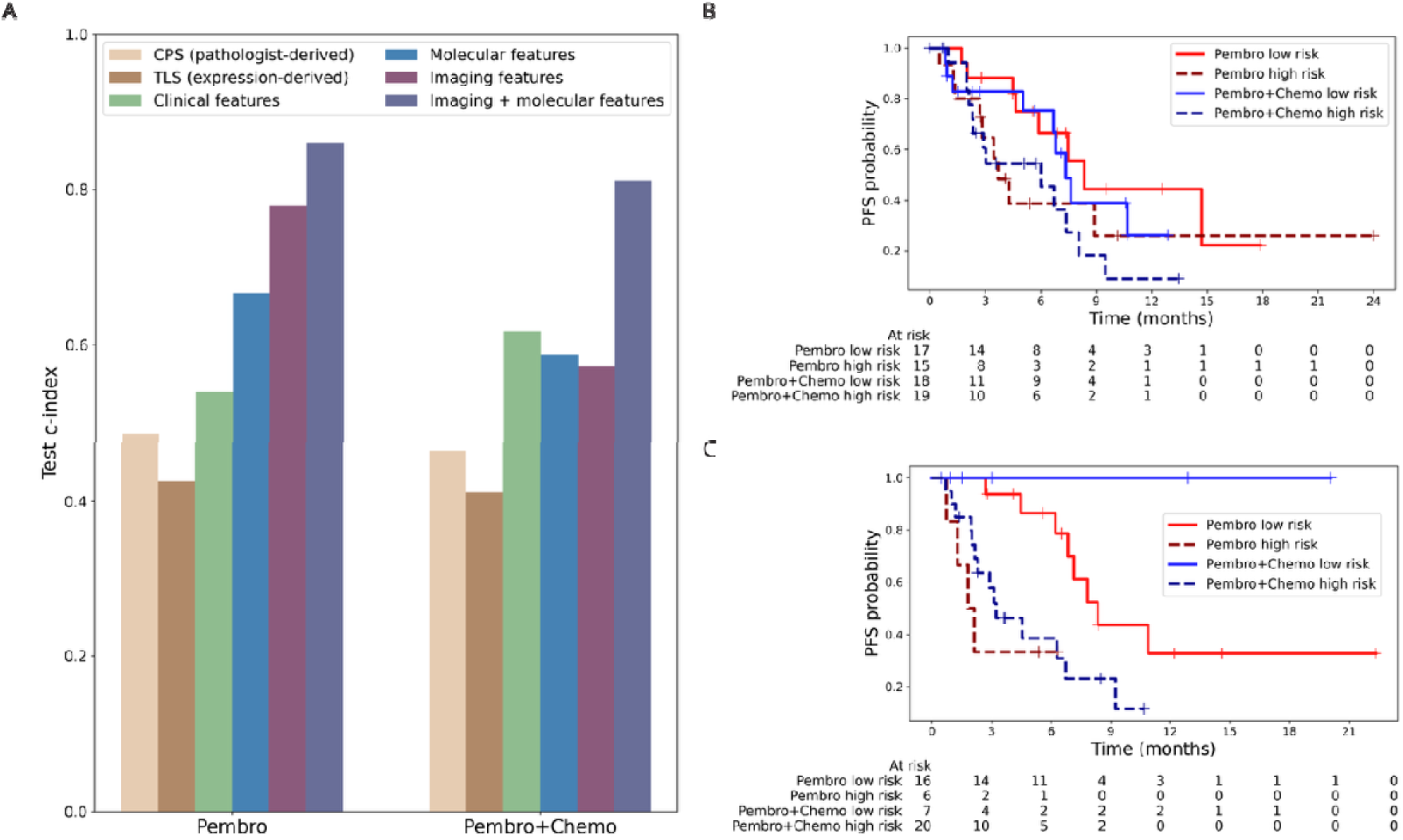
Multimodal risk prediction models outperform individual biomarkers for patient stratification. **(A)** Concordance index comparison across feature types shows imaging and molecular features combined achieve highest predictive performance (c-index: 0.86 Pembro, 0.81 Pembro+Chemo). **(B)** Kaplan-Meier curves for molecular risk signature show trends but lack statistical significance. **(C)** Combined imaging and molecular risk signature achieves significant separation between risk groups in both treatment cohorts (Pembro p=0.004, Pembro+Chemo p=0.028).

Kaplan-Meier survival analysis was performed to evaluate the prognostic efficacy of our molecular risk signature on test patients across both treatment cohorts (Figure 3B). The molecular risk signature demonstrated risk stratification of patient outcomes in both treatment groups, though statistical significance was not achieved in either cohort. In the Pembro cohort, patients classified as low-risk (n=17) showed a trend toward better PFS outcomes compared to those classified as high-risk (n=15), with a log-rank p-value of 0.147. In the Pembro+Chemo cohort, low-risk patients (n=18) also demonstrated a trend toward improved survival compared to high-risk patients (n=19), though the difference was not statistically significant (P = 0.177). As the molecular risk signatures largely consist of genes previously unknown to be related to HNSCC, we explored how the computed scores correlated with known immune-related signatures. In Supplementary Figure S4, we show that the genes in this signature capture both established biology (positively correlating with immune activation signatures and negatively with oncogenic pathways) as well as novel predictive features, providing complementary information that extends beyond individual literature signatures. The molecular risk signature shows consistent directional effects favoring low-risk patients in both treatment settings, however the lack of statistical significance in the test cohort suggests the need for more predictive features to capture the heterogeneity of treatment outcomes in these cohorts.

Kaplan-Meier survival analysis of our combined imaging and molecular risk signature showed clear separation between risk groups in both treatment cohorts (Figure 3C). This multi-modal risk model showed clear and robust separation between patient groups, with low-risk patients having significantly better PFS than high-risk patients in both treatment arms. In the Pembro cohort, low-risk patients (n=16) had much better outcomes than high-risk patients (n=6), with a log-rank p-value of 0.004. In the Pembro+Chemo cohort, low-risk patients (n=7) also had better survival than high-risk patients (n=20), with a log-rank p-value of 0.028. The combined imaging and molecular risk signature performed better than molecular and imaging features alone (see Supplementary Figure S5), with stronger statistical significance and higher c-index values, highlighting that adding imaging data significantly improved patient risk prediction.

### Interpretation of histopathology and molecular risk score

To interpret the model’s predictions, we visualized the learned attention weights by overlaying them on the original H&E images. The heatmaps highlight the tissue regions most predictive of patient outcome (in red). In Figure 4, we provide examples for patients treated with Pembro who were predicted to have different risk levels by the model. These patients had the most extreme predicted risk scores. In the top WSI (low risk patient), we observe that the model pays strong attention (red) to specific regions of interest, which we hypothesize represent TLS clusters. The presence and identification of these clusters could potentially contribute to the low-risk prediction. In the bottom WSI (high risk patient), we see that the model pays stronger attention to regions we hypothesize are necrosis, which are known predictive indicators of poorer patient outcome. Therefore, these necrotic regions may contribute to the high-risk prediction. We provide additional hypothesized annotations for the highest and lowest predicted risk patients in both treatment groups in Supplementary Table S7. We also examined the correlation of the imaging & molecular risk score with our manually curated set of immune-related signatures in Supplementary Figure S6, further supporting the trends we observed.

**Figure 4.**
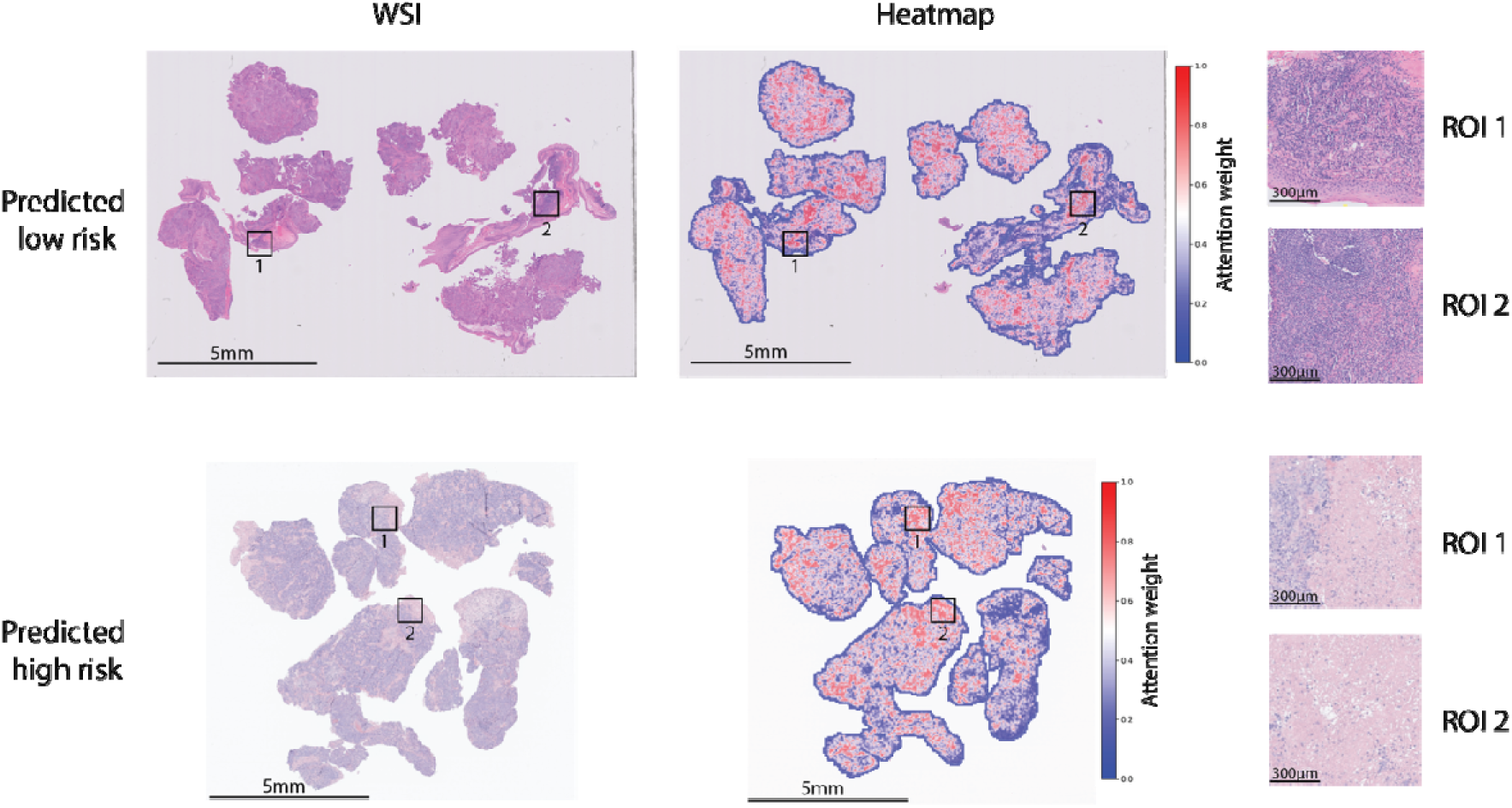
Attention heatmap analysis of WSI using weights learned by imaging & molecular features model. Red corresponds to higher attention areas. Top WSI is from a patient treated with Pembro who was predicted low risk by the model. The model pays stronger attention to TLS regions, as indicated by ROIs, which likely contributes to low-risk prediction. Bottom WSI is from a patient treated with Pembro who was predicted high risk by the model. The model pays stronger attention to necrosis regions as shown in ROIs, which are known predictive indicators of poorer prognosis.

## Discussions and Conclusion

In this real-world study of patients with R/M HNSCC treated with Pembro alone or in combination with Chemo, we performed an integrated analysis leveraging bulk transcriptomic data, digital histopathology, and clinical features to evaluate predictive biomarkers of treatment response. Despite the widespread use of PD-L1 CPS as a biomarker for immunotherapy eligibility, our findings confirm that CPS alone lacks robust prognostic value in R/M HNSCC. This observation is consistent with prior real-world studies showing variable CPS performance outside clinical trial settings, where patient populations are more heterogeneous and are often receiving Pembro in combination with chemotherapy [12, 14]. The subjective nature of CPS scoring by pathologists may further contribute to its limited predictive utility, underscoring the need for more comprehensive biomarkers that better reflect the tumor-immune microenvironment. Availability of PD-L1 CPS across entire RW cohorts, as evidenced by the number of patients with no evaluable CPS measurements in this study, also presents challenges in utilizing the biomarker consistently in this disease setting.

Through systematic modeling of immune-related molecular features, we identified several gene expression signatures significantly associated with PFS in both treatment groups, including those related to cytotoxic T cells, regulatory T cells, and TLS [16]. Notably, subsets of gene signatures were uniquely predictive in either the Pembro monotherapy or combination therapy groups. For example, TLS features enriched for memory T cells, regulatory T cells, and interferon gamma responsive genes typically associated with cytoxic T cell activity [29] were selectively prognostic in the Pembro monotherapy cohort, whereas signatures related to plasma cells, TH2 cells, and NKT cells demonstrated significance only in the Pembro-chemo group. Treatment with either Pembro monotherapy or Pembro plus chemotherapy is generally dictated by PD-L1 CPS status alone. TLS and B cell signatures have been previously associated with improved prognosis in HNSCC [33], and have been associated with favorable response to immunotherapy in multiple indications [18]. The significance of plasma cell and NKT cells in Pembro plus chemo treated patients also points to the importance of immune infiltrate beyond T cells being important for response to an immunotherapy combination regimen, even where PD-L1 CPS is low. These results highlight additional biomarkers which potentially discern treatment benefit of Pembro with chemotherapy and Pembro alone in addition to levels of PD-L1.

Based on emerging studies demonstrating strong associations between TLS-related signatures and clinical outcomes [33], we focused on TLS as a biologically relevant biomarker. Using unsupervised clustering of multiple TLS gene signatures, patients classified as TLS-high exhibited significantly improved PFS, particularly in the Pembro monotherapy group. Moreover, histologically defined TLS presence on H&E-stained WSIs showed strong concordance with transcriptomic TLS scores with five out of six signatures demonstrating significant correlations with morphological assessments. This consistency strengthens the utility of TLS as a predictive biomarker.

To build upon these literature-derived biomarkers, we developed treatment-specific molecular signatures that integrate both established immune pathways and novel genes, achieving improved risk stratification through this additive approach that captures biology not represented in any single curated signature. Our systematic evaluation revealed a clear performance hierarchy: traditional pathological markers (CPS, TLS signature) showed limited discriminative accuracy (c-index 0.43-0.49), clinical features performed modestly (0.54-0.61), individual molecular signatures achieved moderate performance (0.59-0.66), while PLIP-derived imaging features demonstrated superior capability (0.57-0.78). The combination of imaging and molecular features achieved highest performance with c-index values of 0.86 and 0.81 in pembrolizumab and combination therapy cohorts, respectively, alongside statistically significant risk stratification in both groups. This superior performance reflects the complementary nature of the multimodal data: transcriptomic signatures capture the overall immune landscape and activation states, while imaging data provides crucial spatial organization and morphological context not available in bulk RNA sequencing. This spatial context appears critical for patient-specific risk assessment, as the tumor microenvironment’s architectural features, such as TLS organization, necrotic regions, and cellular infiltration patterns, vary substantially between patients and may modulate the functional significance of molecular signatures.

These findings have important implications for clinical implementation. In routine clinical practice, many patients may lack one or more of these modalities, particularly RNA sequencing data, due to cost, tissue availability, or time constraints. Our results suggest a practical framework for such real-world scenarios: when RNA is available, the multimodal approach (digital pathology features + molecular signatures) provides optimal risk stratification. When RNA is not available, digital pathology features alone demonstrate superior performance compared to CPS or clinical features alone, suggesting that computational analysis of routine H&E slides can provide meaningful prognostic information without requiring additional molecular testing. For patients identified as high-risk based on imaging features alone, physicians may consider whether the added benefit of molecular profiling justifies the additional testing burden. In resource-limited settings, prioritizing imaging-based analysis may provide the most effective approach to improve patient stratification beyond standard pathological assessment.

This study had some limitations which should be considered. The sample sizes of the RW treatment cohorts used limited the sizes possible for test sets for our multimodal models. The availability of PD-L1 CPS data across the RW cohort was also incomplete. The ideal endpoint to assess for associations of biomarkers with outcomes would be OS, which is difficult to assess in RW cohorts due to immortal time bias when determining survival with respect to treatment timings and disease diagnoses [34].

Additionally, our analysis was limited to patients with both high-quality transcriptomic and histopathological data, which may not represent the broader R/M HNSCC population typically seen in clinical practice. Future work should focus on prospective validation in larger independent cohorts, systematic evaluation of model performance with different combinations of available data modalities, and exploration of integration into clinical decision-making algorithms that account for varying levels of biomarker availability across different healthcare settings.

In summary, this study demonstrates that integrated molecular and spatial biomarkers outperform single-modality predictors such as PD-L1 CPS in stratifying R/M HNSCC patients for immunotherapy response. TLS features offer strong and consistent prognostic value, and their integration with imaging data enhances risk stratification. Importantly, when comprehensive molecular profiling is not available, computational pathology features from routine H&E slides provide superior prognostic information compared to traditional pathological markers alone, offering a practical pathway for improved patient stratification in real-world clinical settings. The multimodal approach captures complementary tumor biology aspects that individual biomarkers cannot adequately represent, providing a foundation for more precise personalized treatment strategies in R/M HNSCC immunotherapy.

## Data availability

De-identified, individual-level data used in this research were collected in a real-world healthcare setting by Tempus AI, Inc. and are subject to controlled access for privacy and contractual reasons. The ethics committee and/or informed consent do not allow for public availability. Derived data supporting the conclusions of this article are included within the article and its additional files.

## Authors’ Contributions

**MB** contributed to conceptualization, data curation, formal data analysis, supervision, project administration, and writing of the original draft as well as review and editing of the manuscript. **NT** also contributed to conceptualization, performed formal data analysis, and was responsible for writing the original draft as well as reviewing and editing the manuscript. **IK** was involved in data curation and manuscript review and editing. **LKB, JZ**, and **SS** contributed to conceptualization, supervision, and manuscript review and editing. **YS** and **AM** performed annotation and identification of TLS markers from H&E data. **MG**, **DS**, and **HS** contributed to manuscript review and editing.

## Acknowledgments

The authors acknowledge the use of ChatGPT (OpenAI) for initial language editing and refinement of the manuscript. All subsequent revisions and final editing were performed by the authors.

## Supplementary Methods

### Prediction of HPV status

HPV status was only known for 55 patients from lab tests (35 positive and 20 negative patients), so we imputed HPV status for the remaining patients. We used CDKN2A expression as a proxy for HPV positivity [35, 36]. We split the 55 patients into a train and test set (70/30 split), and using a logistic regression model we predicted binary HPV status from expression level on the training patients. On the testing patients, we achieved an 88% testing accuracy. Finally, using the trained model, we imputed HPV for the remaining patients; in total, 124 patients were marked as HPV positive and 106 as HPV negative.

### Clinical model

Clinical information included: age, PD-L1 CPS score (annotated by pathologists), gender, biopsy location and HPV status. We did not include ECOG due to high missingness of values. Using these clinical attributes, we trained four treatment-specific models to predict PFS outcomes. We evaluated the following models: Cox’s proportional hazard (coxph), Cox’s proportional hazard with elastic net penalty (coxnet), random survival forest, and XGBoost [37, 38]. For each model, we subset patients based on the first line of therapy treatment (Pembro monotherapy or Pembro+Chemo regimen) and evaluated the model’s performance using c-index. We provide the comparative results of this analysis in Supplementary Figure S7.

### Pathology feature extractor evaluation

Several pathology WSI encoders were evaluated to determine the best performing model. We evaluated the following feature extractors: PLIP [25], UNI2 [39], H-optimus-0 [40], H-optimus-0 [40], and Virchow2 [41] to determine which feature extraction embeddings were most informative in predicting PFS and stratifying risk. In both first-line therapy treatment settings, the embeddings from PLIP led to the highest test c-index (see Supplementary Figure S8).

Authors contribution (subject to change):

Conceptualization: Merzu, Sriram, Nitya, Lauren

Data curation: Merzu, Indu

Formal analysis: Merzu, Nitya, Indu

Supervision: Merzu, Lauren, Sri, James and Brandon

Writing – original draft: Merzu, Nitya

Writing – review & editing: All authors

