## Supplemental Figures & Tables for "Integrated Histopathology–Transcriptomic Biomarker Enhances Survival Prediction in HNSCC Patients Treated with Immunotherapy"

Building 2, 777 Scudders Mill Rd, Princeton, NJ 08540

Authors contribution (subject to change):

Conceptualization: Merzu, Sriram, Nitya, Lauren

Data curation: Merzu, Indu

Formal analysis: Merzu, Nitya, Indu

Supervision: Merzu, Lauren, Sri, James and Brandon

Writing – original draft: Merzu, Nitya

Writing – review & editing: All authors

Keywords: Head and neck cancer, tertiary lymphoid structures, clinical outcomes, treatment

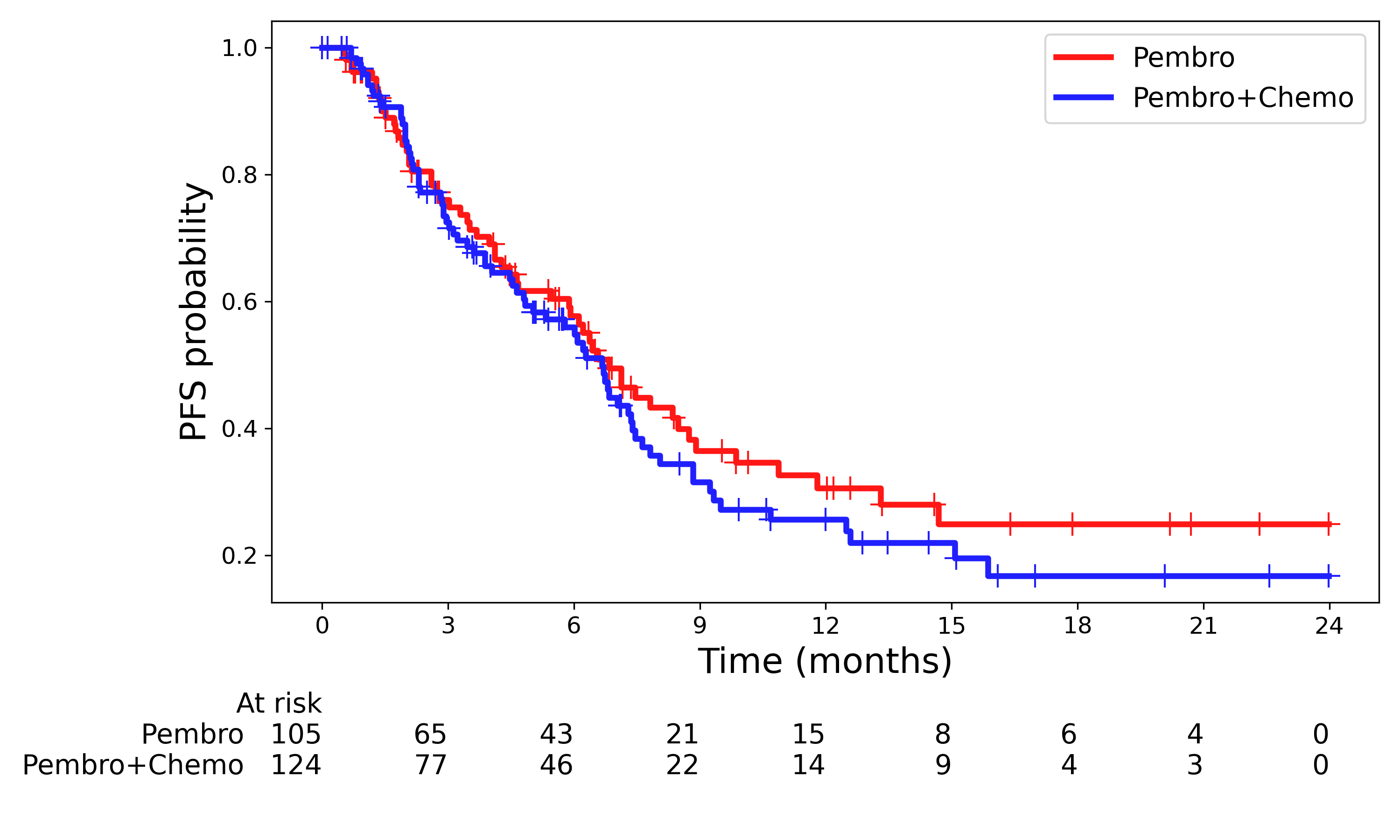

Supplementary Figure S1: KM plots for PFS for the treatment regimens. No significant PFS advantage between the treatment groups

| ***Treatment***  ***Groups*** | ***TLS (B- & T- ) cells receptors*** | ***TLS (mature DC cells)*** | ***TLS core genes*** | ***TLS(chemokine)*** | ***TLS (tfh) cell*** | ***TLS (Th1- & B-) cell*** | ***model*** |
| --- | --- | --- | --- | --- | --- | --- | --- |
| ***Pembro+Chemo*** | 0.491 | 0.83 | 0.001 | 0.051 | 0.191 | 0.469 | KM |
| ***Pembro*** | 0.242 | 0.11 | 0.07 | 0.038 | 0.041 | 0.002 | KM |
| ***Pembro+Chemo*** | 0.183 | 0.491 | 0 | 0.045 | 0.235 | 0.315 | Cox |
| ***Pembro*** | 0.217 | 0.087 | 0.09 | 0.038 | 0.031 | 0.004 | Cox |

Supplementary Table S1: KM & Cox model for TLS where the numbers are the p-value from the two models. Majority of the TLS signatures showed statistically significant with better PFS value both in Kaplan-Meier (KM) and Cox Proportional Hazards Model (Cox).

| 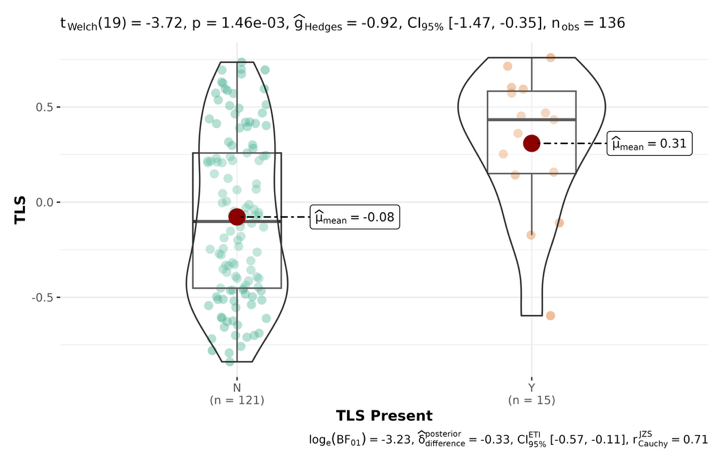 | 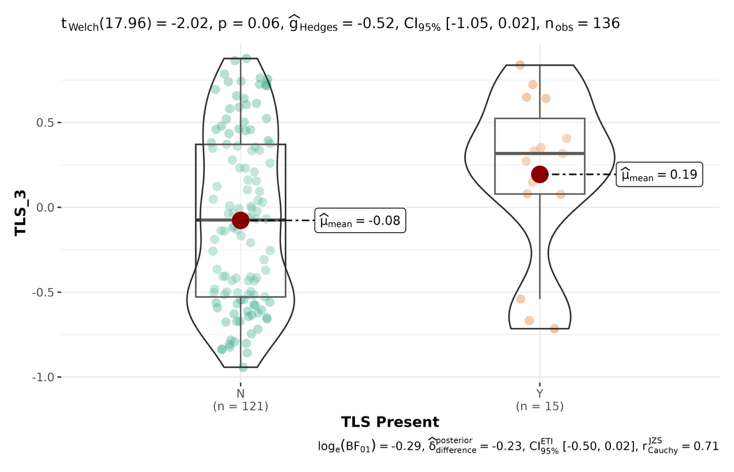 |
| --- | --- |
| 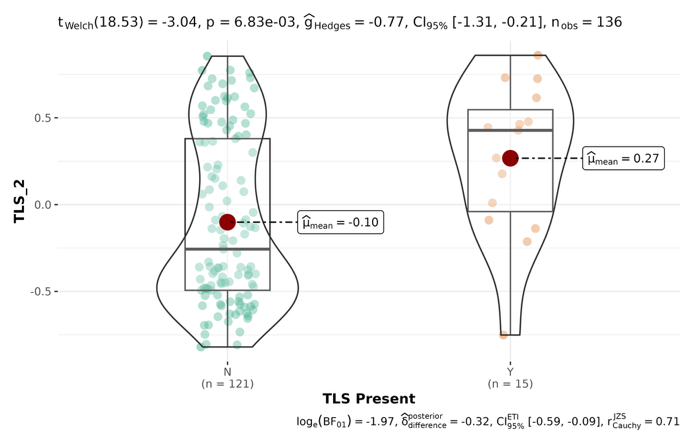 | 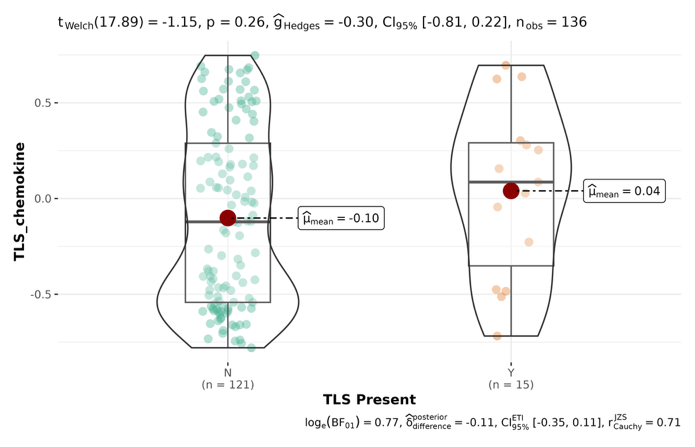 |
| 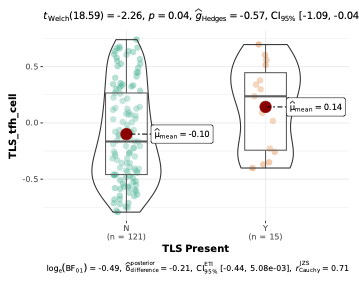 | 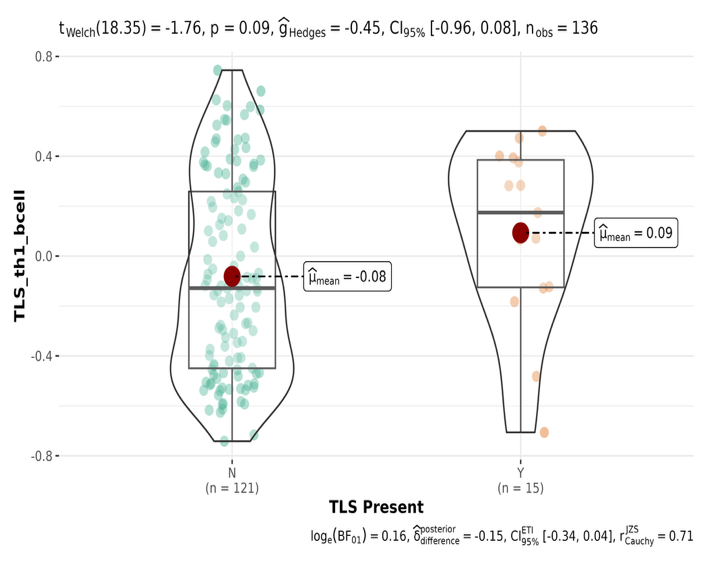 |

Supplementary Figure S2: TLS structure with transcriptome data ; 5/6 signatures have p-value < 0.10 where TLS with chemokines signature has p-value greater than 0.10.

| TLS Present | Kmeans Cluster | | |
| --- | --- | --- | --- |
|  | HIGH | LOW | TOTAL |
| YES | 80% (12) | 20% (3) | 100% (15) |
| NO | 45% (58) | 55% (72) | 100% (130) |
| TOTAL | 48% (70) | 52% (75) | 100% (145) |

Supplementary Table S2: TLS clusters correlated with TLS structure . TLS-positive cases are much more likely to fall into the HIGH cluster (80% observed vs ~48% expected).

| 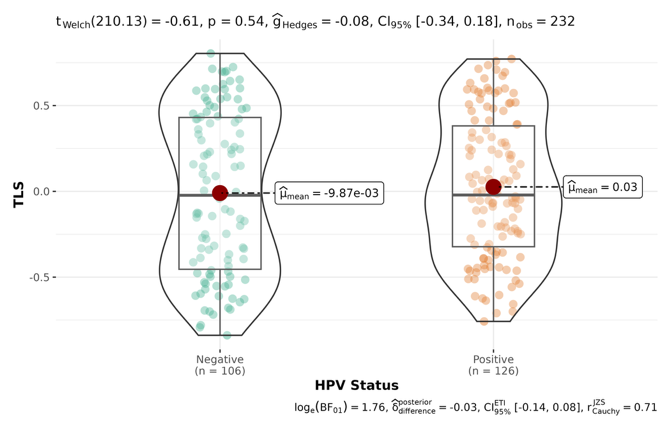 | 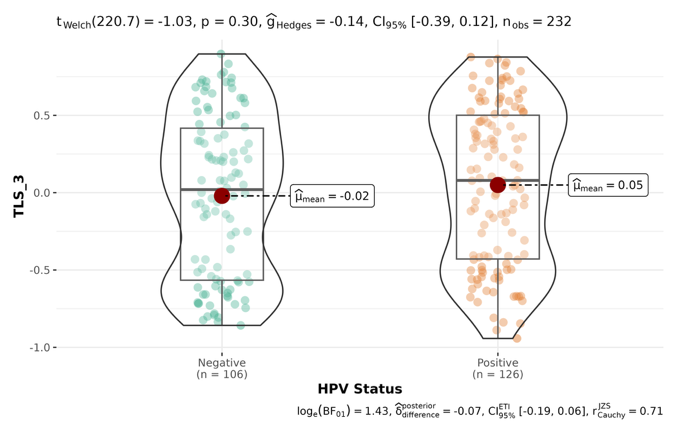 |
| --- | --- |
| 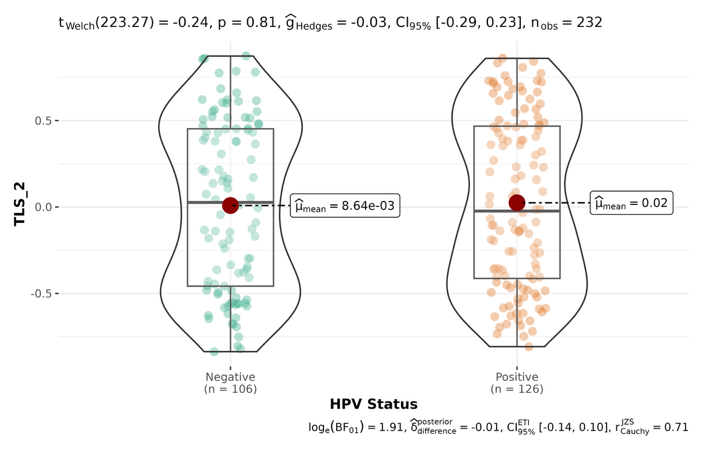 | 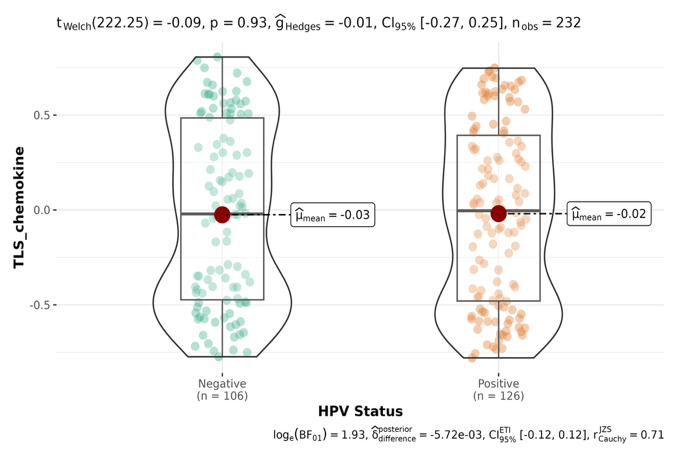 |
| 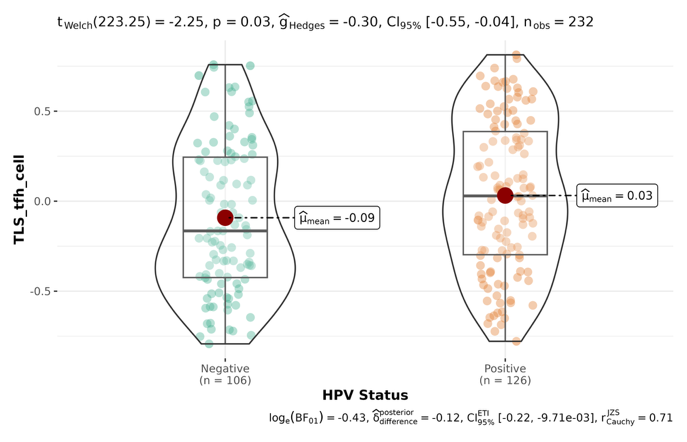 | 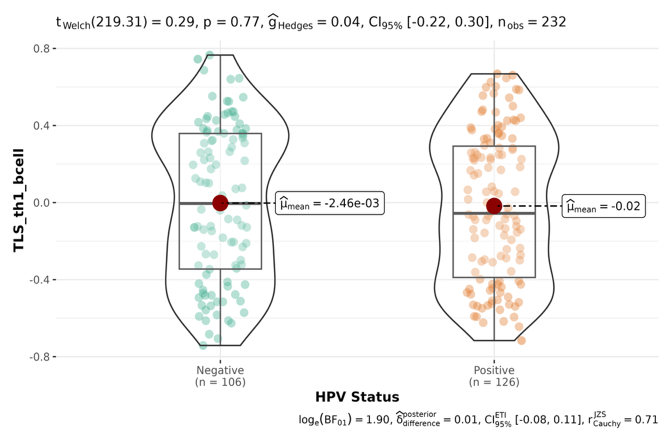 |

Supplementary Figure S3: TLS signature correlation with HPV. Only the T follicular helper (Tfh) cell–associated signature showed a significant correlation with HPV status

| **TLS Present** | **HPV Status** | | |
| --- | --- | --- | --- |
|  | Positive | Negative | TOTAL |
| **YES** | 47%(7) | 53%(8) | 100%(15) |
| **NO** | 59%(71) | 41%(50) | 100%(121) |
| **TOTAL** | 57%(78) | 43%(58) | 100%(136) |

Supplementary Table S3: HPV status and TLS presence, as determined by both H&E-stained WSI and bulk transcriptomic data. No significant association was observed between HPV positivity and TLS presence based on histopathologic evaluation from WSI).

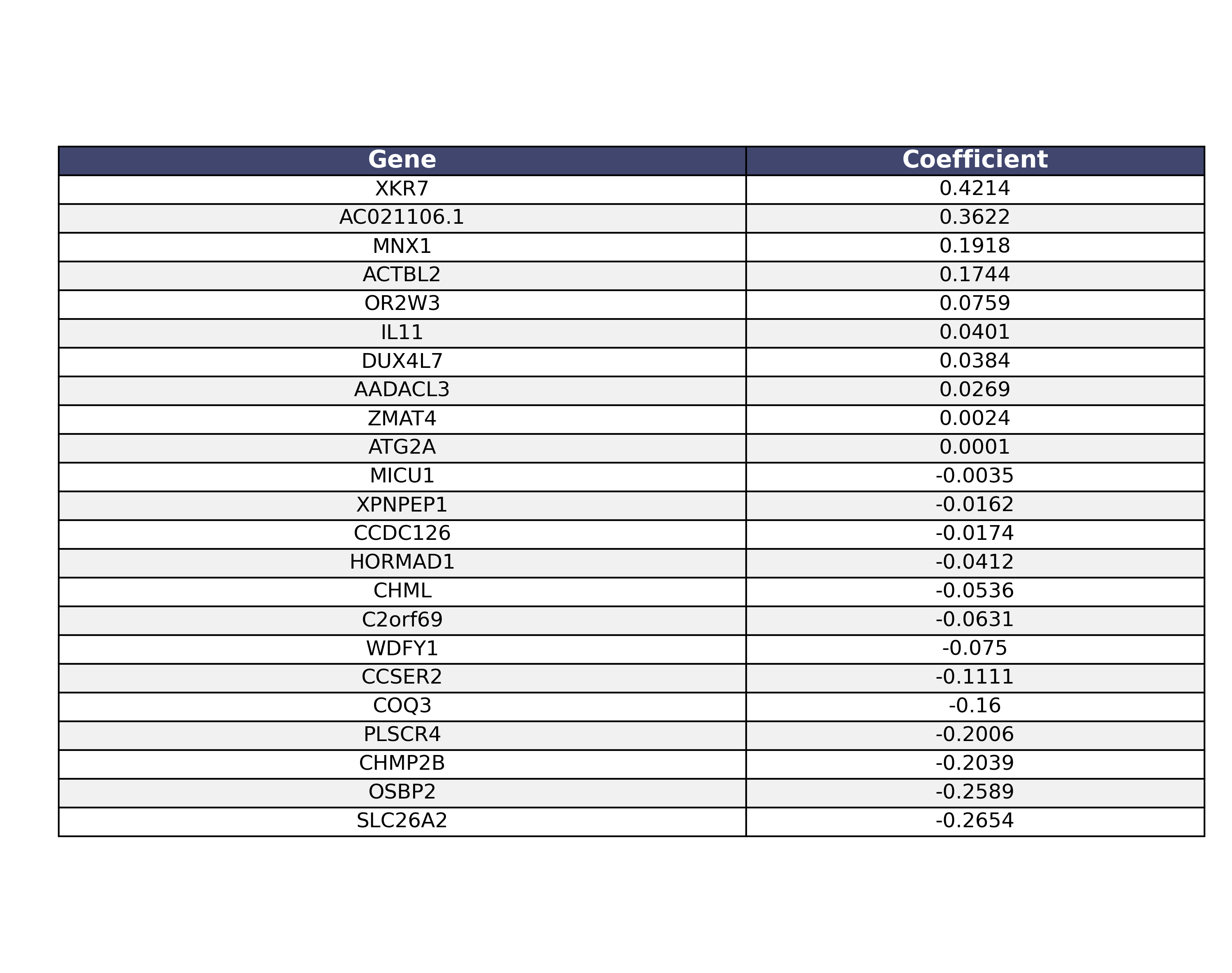

Supplementary Table S4: Here we present the 23 genes and corresponding coefficients that compose the molecular risk prediction model for Pembro patients.

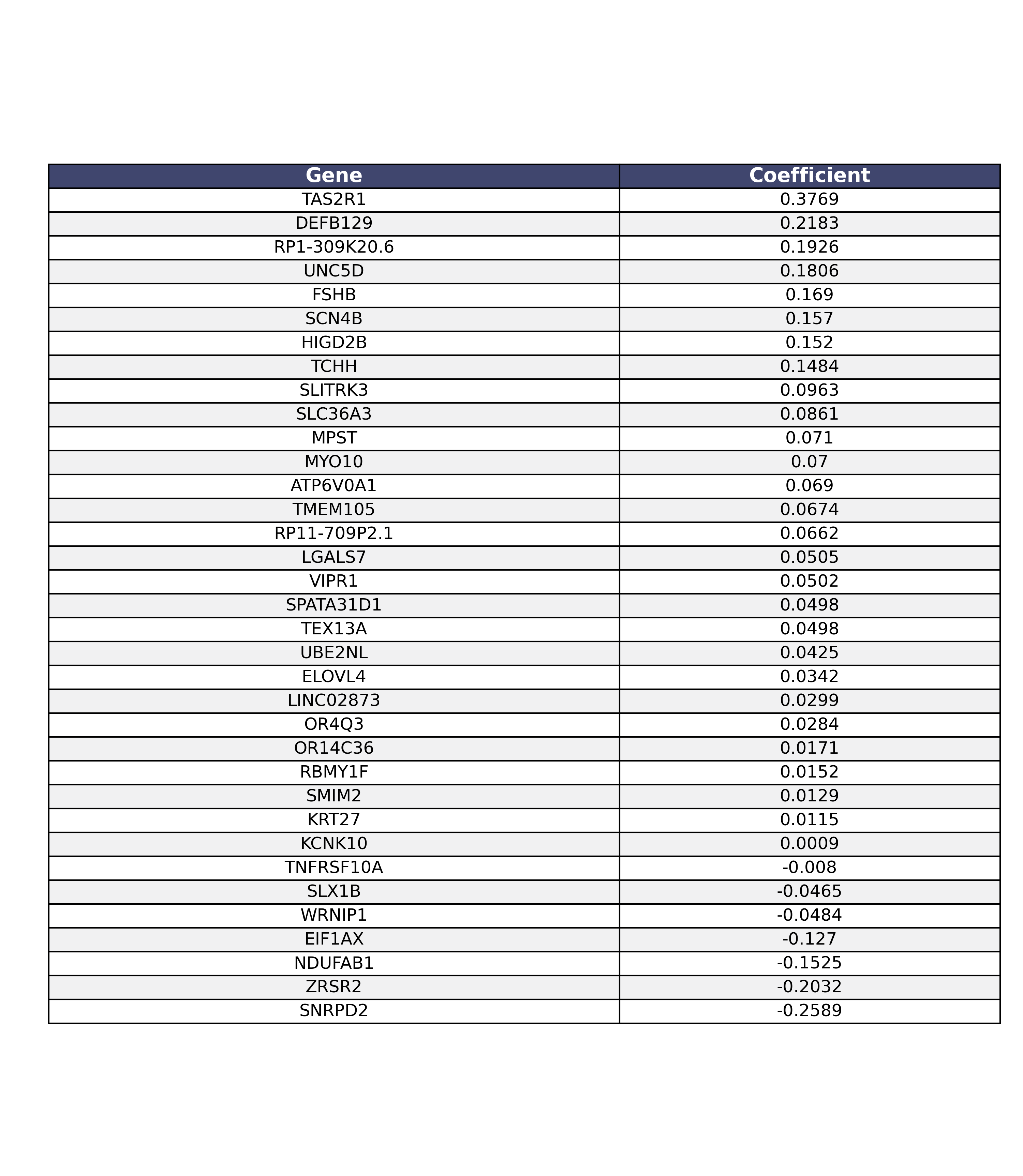

*Supplementary Table S5: Here we present the 35 genes and corresponding coefficients that compose the molecular risk prediction model for Pembro+Chemo patients.*

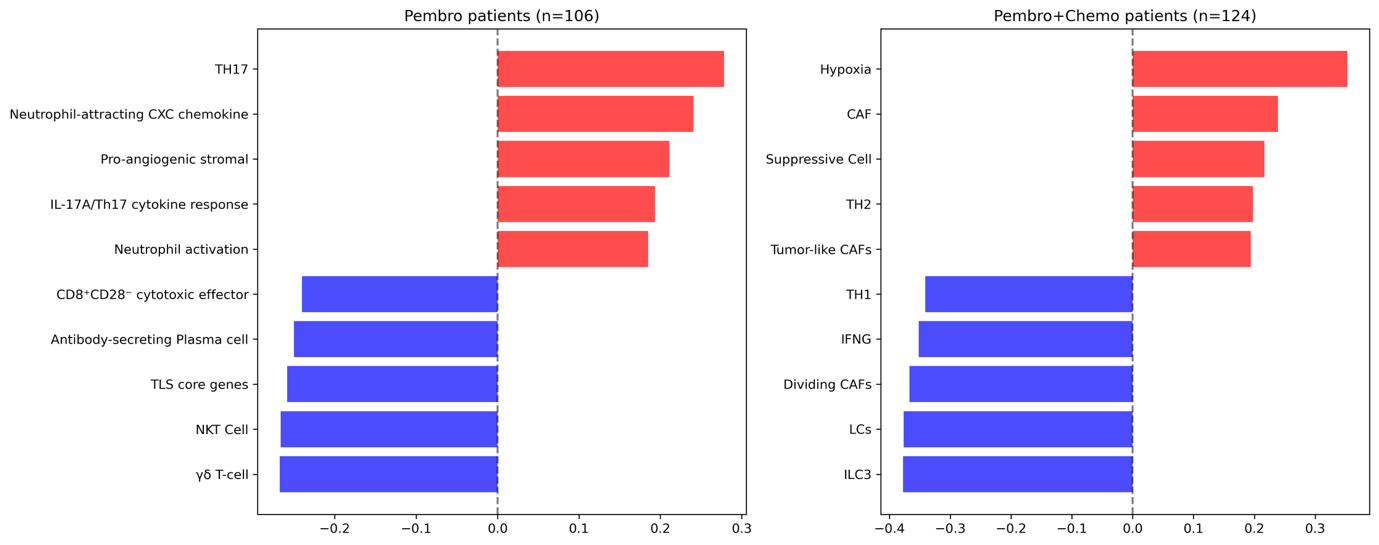

*Supplementary Figure S4: Correlation of molecular risk score with manually curated set of immune-related signatures. We show the top 5 positive and negative correlation signatures. We find for both treatment groups, higher predicted risk is correlated with pro-tumor molecular signatures, and lower predicted risk is correlated with anti-tumor molecular signatures.*

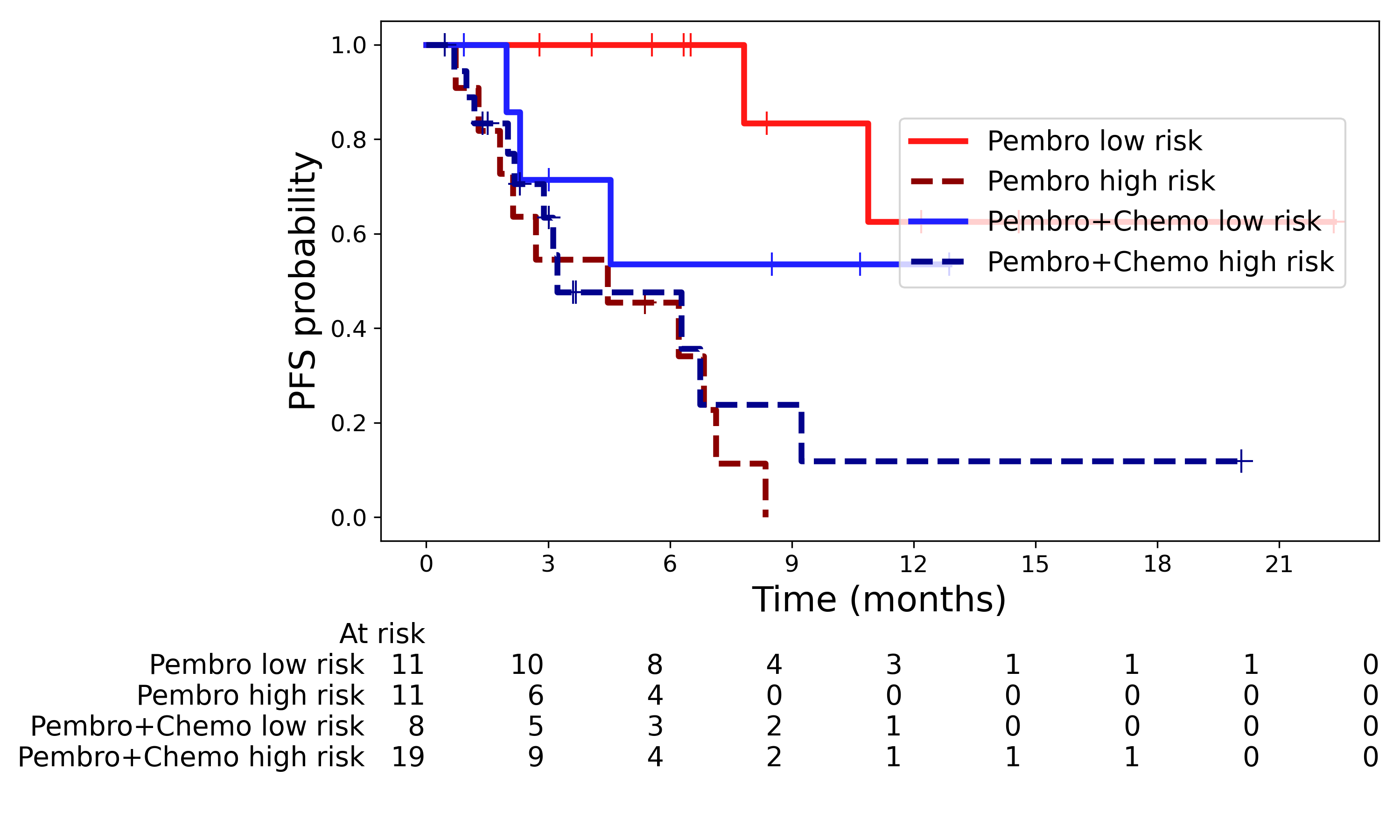

*Supplementary Figure S5: Kaplan-Meier curves for imaging risk model show trends but lack statistical significance for Pembro+Combo group (p=0.22). The stratification was significant for Pembro test patients (p=6.5e-5).*

| Model type | Hidden dimension | Dropout rate | Learning rate |
| --- | --- | --- | --- |
| Imaging features | 224 | 0.2 | 8e-4 |
| Imaging and molecular features | 160 | 0.3 | 5e-4 |

*Supplementary Table S6: When training the imaging-based models, we performed a hyperparameter sweep. Here, we report the hyperparameters that led to the best test c-index and risk stratification on test patients for imaging & imaging and molecular models. The hidden dimension refers to the dimension the image feature embeddings were projected to before input into the MIL model.*

| Treatment | Predicted risk | Hypothesized ROI subtype | Treatment | Predicted risk | Hypothesized ROI subtype |
| --- | --- | --- | --- | --- | --- |
| Pembro | High | Large area of necrosis | Pembro | Low | TLS, lymphocyte infiltration into tumor nest |
| Pembro | High | Keratinized tumor nests | Pembro | Low | Solid tumor nests with high density of ICs |
| Pembro+Chemo | High | Tumor nests with apoptotic cells | Pembro+Chemo | Low | TLS, IC infiltration around tumor nest |
| Pembro+Chemo | High | Keratinized tumor nests | Pembro+Chemo | Low | Highly inflamed stroma with high density of ICs and infiltration of tumor cells |

*Supplementary Table S7: We present hypothesized characterizations for select regions of interest (ROIs) for Pembro and Pembro+Chemo patients. The chosen patients represent the ones with the highest and lowest predicted risk scores. The ROIs chosen are regions considered high attention by the imaging model. From this sample of images we analyzed, we hypothesize that across both treatment groups the model pays high attention to biologically relevant features. In patients predicted high-risk, we propose that regions of necrosis and tumor nests are driving the model’s prediction. For patients predicted low-risk, we propose that regions of TLS and immune cell (IC) infiltration are driving the model’s prediction.*

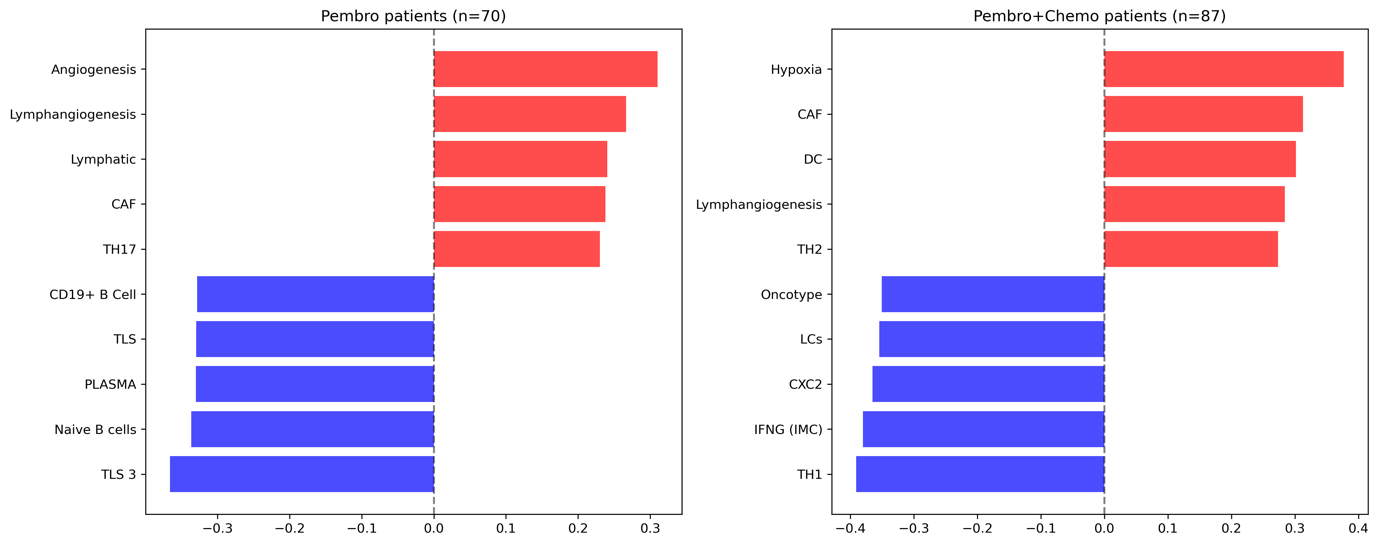
*Supplementary Figure S6: Correlation of imaging & molecular risk score with manually curated set of immune-related signatures. We show the top 5 positive and negative correlation signatures. We find for both treatment groups, higher predicted risk is correlated with pro-tumor molecular signatures, and lower predicted risk is correlated with anti-tumor molecular signatures.*

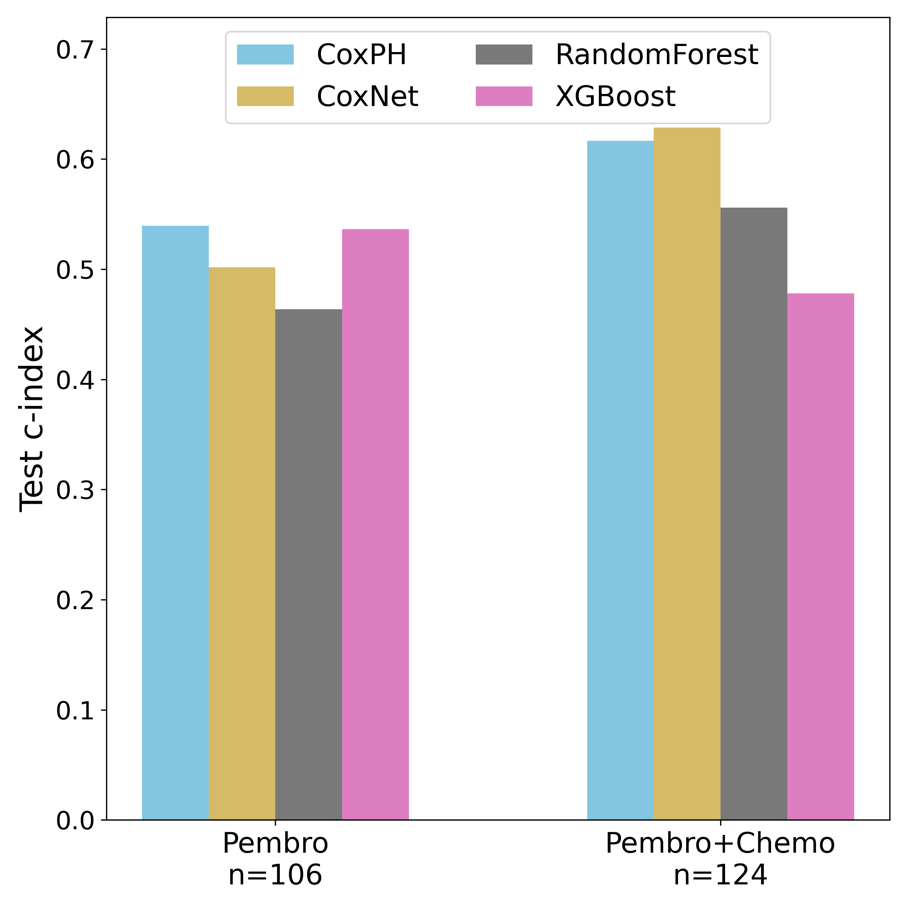

*Supplementary Figure S7: PFS C-index on test patients using clinical features across different predictive models. All models showed similar performance, and for comparison to other modalities we chose the CoxPH model*

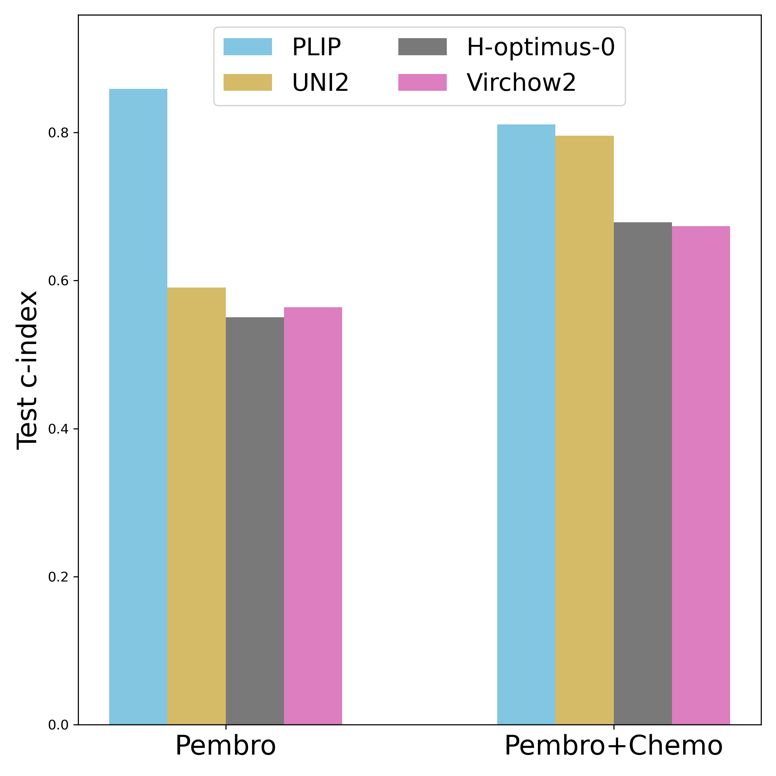

*Supplementary Figure S8: To create the imaging features, we tested four state of the art (SOTA) pathology foundation models. Here, we present the results of the different feature embeddings in downstream PFS outcome prediction when combining imaging and molecular features. We found that PLIP consistently outperformed the other models, especially in the Pembro-only setting.*
